# Monogenic Syndromes as a Cause of Adverse Drug Reactions in the Russian Population

**DOI:** 10.64898/2026.02.13.26346297

**Authors:** Anastasiia A. Buianova, Valery V. Cheranev, Anna O. Shmitko, Iuliia A. Vasiliadis, Galit A. Ilyina, Oleg N. Suchalko, Mikhail Iu. Kuznetsov, Vera A. Belova, Dmitriy O. Korostin

## Abstract

**Introduction:** Adverse drug reactions (ADRs) remain a major public health issue, and genetic factors contribute importantly to interindividual variability in drug response. Pharmacogenetic testing helps reduce ADR risk by optimizing drug selection and dosage, particularly in monogenic disorders.

**Material and Methods:** Whole-exome sequencing of 6,739 samples from the Russian population was performed using the MGIEasy Universal DNA Library Prep Set on the DNBSEQ-G400 platform (MGI). Variants in 48 genes were examined, focusing on inherited arrhythmias (Long QT syndrome, Short QT syndrome, Timothy syndrome, Andersen-Tawil syndrome, Brugada syndrome, Atrial fibrillation, Catecholaminergic polymorphic ventricular tachycardia), enzyme deficiencies (Glucose-6-Phosphate Dehydrogenase Deficiency [G6PDD], Porphyrias), Dravet Syndrome (DS) and Malignant Hyperthermia (MH). All identified variants had been reported at least once as pathogenic (P) or likely pathogenic (LP) in ClinVar, along with those occasionally classified as variants of uncertain significance (VUS). Each variant was manually re-evaluated according to ACMG criteria.

**Results:** A total of 75 unique variants in 18 genes were observed in 119 individuals (1.77%), including 21 carriers and 13 women with a *G6PD* mutation. Of these, 46 variants were classified as P, 21 as LP, and 8 as VUS. Missense variants accounted for the largest proportion (73.33%). The most affected genes were *KCNQ1* (24/119), which exhibited the highest number of unique variants (18), *G6PD* (20/119), *SCN1A* (15/119), and *RYR1* (14/119). Regarding associated conditions, mutations linked to arrhythmias were found in 51 individuals, MH in 27, G6PDD in 20, DS in 15, and Porphyrias in 6.

**Conclusions:** Incorporating genetic information on both common and rare clinically actionable variants into therapeutic decision-making has the potential to improve medication safety, reduce preventable ADRs, and enhance the effectiveness of personalized pharmacotherapy.

## Introduction

Adverse drug reaction (ADR) is defined as a harmful and unintended response to a medication occurring at standard doses with an established causal relationship [1]. Although the risk of ADRs in elderly patients was reported to be four times higher than in younger individuals (16.6% vs. 4.1%), the proportion of preventable cases was 88% and 24%, respectively, in a 2002 meta-analysis [2]. A subsequent analysis of 5,707 emergency hospitalizations, conducted twelve years later, indicated that 5% of cases were directly attributable to ADRs, with advanced age and the expected polypharmacy remaining the primary risk factors [3]. However, hospital-based ADR statistics differ: among 5,644 cases studied from 2020 to 2023 in a Chinese hospital, severe ADRs (*n* = 408, 7.2%) were more common in middle-aged patients (46–65 years, 36.77%), predominantly female (two-thirds), frequently associated with intravenous administration (53.92%), and primarily affecting the circulatory system (53.19%) [4]. According to VigiBase data (2010–2019), fatal ADRs accounted for 1.3% (43,685 cases) of all reported reactions, with antineoplastic and immunomodulatory drugs (denosumab, lenalidomide, thalidomide) representing the leading cause [5]. Between Q3 2014 and Q3 2024, approximately 2.7 million ADR reports were registered in the FAERS database, with each report linking an event to a single drug [6]. Given that the FDA approved roughly 494 drugs during this period, the increase in FAERS reports is likely more reflective of changes in pharmacovigilance systems and awareness than of actual incidence [7].

Genetics plays a central role in ADR susceptibility, as UK Biobank data demonstrate that nearly all individuals (99.5%) carry at least one pharmacogenetic variant capable of eliciting an atypical drug response, and 24% of participants had already been exposed to the corresponding medications [8]. While widely implemented pharmacogenetic testing programs (e.g., PharmCAT [9]) focus on common polymorphisms (allele frequency >3%) to optimize dosing of frequently used medications or guide therapeutic substitutions, the clinical significance of rare variants associated with monogenic syndromes remains insufficiently explored. The presence of such variants implies a life-threatening risk of severe idiosyncratic reactions upon exposure to specific medications during essential treatment courses [10]. ADRs can manifest for the first time in advanced age, as illustrated by a case of malignant hyperthermia (MH) in a 79-year-old woman following succinylcholine administration in the absence of prior genetic testing [11].

With the increasing accessibility of whole-exome sequencing (WES) in the Russian Federation, it has become feasible to construct a population-level genetic risk profile for severe ADRs, which is particularly relevant for a country with pronounced ethnic diversity. Integration of such data into preoperative screening, targeted genetic testing, and clinical protocols could substantially reduce the risk of fatal outcomes.

## Materials and Methods

### 2.1. Sample collection

The study included 6,739 anonymized whole-exome sequencing datasets generated between 2020 and 2025. The cohort represents an unselected population-based sample, including neonates, pediatric and adult patients, as well as individuals undergoing carrier screening or preconception testing. Samples were primarily collected in Moscow hospitals, with some patients referred from other regions. Available metadata included only sex and the type of exome enrichment kit. Although clinical and phenotypic information existed for a subset of samples, these data were not incorporated, as the analysis was conducted using anonymized exome data exclusively.

### 2.2. DNA Sequencing and Bioinformatic Data Processing

DNA libraries were prepared from 500 ng of genomic DNA from peripheral blood samples using the MGIEasy Universal DNA Library Prep Set (MGI Tech, Shenzhen, China). DNA fragmentation was performed via ultrasonication (Covaris S-220, Covaris, Inc., Woburn, MA, USA) to an average fragment size of 250 bp. Exome enrichment was conducted using Agilent SureSelect Human All Exon v6/v7/v8 probes [12], and library concentrations were measured with Qubit Flex using the dsDNA HS Assay Kit (Invitrogen, Waltham, MA, USA). Library quality and fragment size distribution were assessed with Bioanalyzer 2100 (Agilent Technologies, Santa Clara, CA, USA). Libraries were circularized and sequenced in paired-end mode (PE100) on the DNBSEQ-G400 platform (MGI Tech), achieving an average coverage of 100×.

Raw sequencing reads (FastQ) were generated using basecallLite (MGI Tech) and quality-checked with FastQC v0.12.1 [13]. Low-quality bases and adapter sequences were trimmed using BBMap v38.96 [14]. Reads were aligned to the human reference genome GRCh38 with bwa-mem2 v2.2.1 [15], and BAM files were processed with SAMtools v1.9 [16]. Duplicate marking and enrichment metrics were obtained using Picard v2.22.4 [17]. Variant calling was performed using bcftools v1.9 [18] and DeepVariant v1.5.0 [19], followed by annotation with VEP v113 [20]. Finally, MultiQC v1.16 [21] was used for comprehensive quality control of the sequencing and analysis outputs.

### 2.3. Clinical Interpretation

Clinical significance of all identified variants, which had been reported at least once as pathogenic (P) or likely pathogenic (LP) in ClinVar, along with variants occasionally classified as of uncertain significance (VUS), was assessed according to ACMG/AMP guidelines [22]. In cases where a single rsID corresponded to multiple alternative substitutions at the same genomic position, all annotated variants were additionally evaluated independently using the ACMG/AMP classification framework. Variants were additionally reviewed manually in IGV [23]. The analysis focused on genes associated with inherited arrhythmia syndromes (including long and short QT syndromes, Brugada syndrome, Timothy syndrome, Andersen–Tawil syndrome, atrial fibrillation, and catecholaminergic polymorphic ventricular tachycardia), MH, glucose-6-phosphate dehydrogenase deficiency (G6PDD), porphyrias, and Dravet syndrome (DS). Gene–disease associations were primarily determined based on data from OMIM [24], with additional validation of the level of evidence for gene–disease relationships using the GenCC database (not lower than the “Limited” level of evidence) [25]. As an exception, the *TRPM4* gene was included because loss-of-function (LOF) variants in this gene have been reported to cause Brugada syndrome [26]. In the interpretation of *RYR1* variants associated with MH, we applied not only the standard ACMG/AMP criteria but also the disease-specific recommendations of the ClinGen Malignant Hyperthermia Susceptibility (MHS) Variant Curation Expert Panel, developed specifically for *RYR1*. These criteria account for gene-specific features of the available evidence, including modified application of the PS1, PM5, and PS4 rules, as well as the high prevalence of variants of uncertain significance [27].

It was taken into account that several genes are involved in the pathogenesis of multiple clinical syndromes, reflecting the overlapping nature of molecular mechanisms and increasing their clinical and pharmacogenetic relevance. The custom gene panel comprised 48 genes: *ABCC9, ALAD, ALAS2, ANK2, CACNA1C, CACNA1S, CACNB2, CALM1, CALM2, CALM3, CASQ2, CAV3, CPOX, FECH, G6PD, GJA5, GPD1L, HCN4, HMBS, KCNA5, KCND3, KCNE1, KCNE2, KCNE3, KCNH2, KCNJ2, KCNJ5, KCNQ1, MYL4, NPPA, NUP155, PKP2, PPOX, RYR1, RYR2, SCN1A, SCN1B, SCN2B, SCN3B, SCN4B, SCN5A, SNTA1, STAC3, TECRL, TRDN, TRPM4, UROD, UROS*. Among these, 32 genes were associated exclusively with autosomal-dominant syndromes under study, 8 exclusively with autosomal-recessive conditions, 6 with both autosomal-dominant and autosomal-recessive inheritance, and 2 genes were located on the X chromosome.

## Results

### 3.1. Overall characteristics of clinically significant variants

In the studied cohort, 119 individuals were identified with clinically significant rare variants associated with monogenic disorders predisposing to ADRs (Figure 1A). Among them, 21 individuals (17.65% of genotype-positive subjects) carried at least one clinically significant rare variant associated with a monogenic disorder conferring pharmacogenetic risk, corresponding to an overall prevalence of 1.77% (Figure 1A). Additionally, 13 women were carriers of variants in the *G6PD* gene (X-linked) (10.92%).

**Figure 1.**
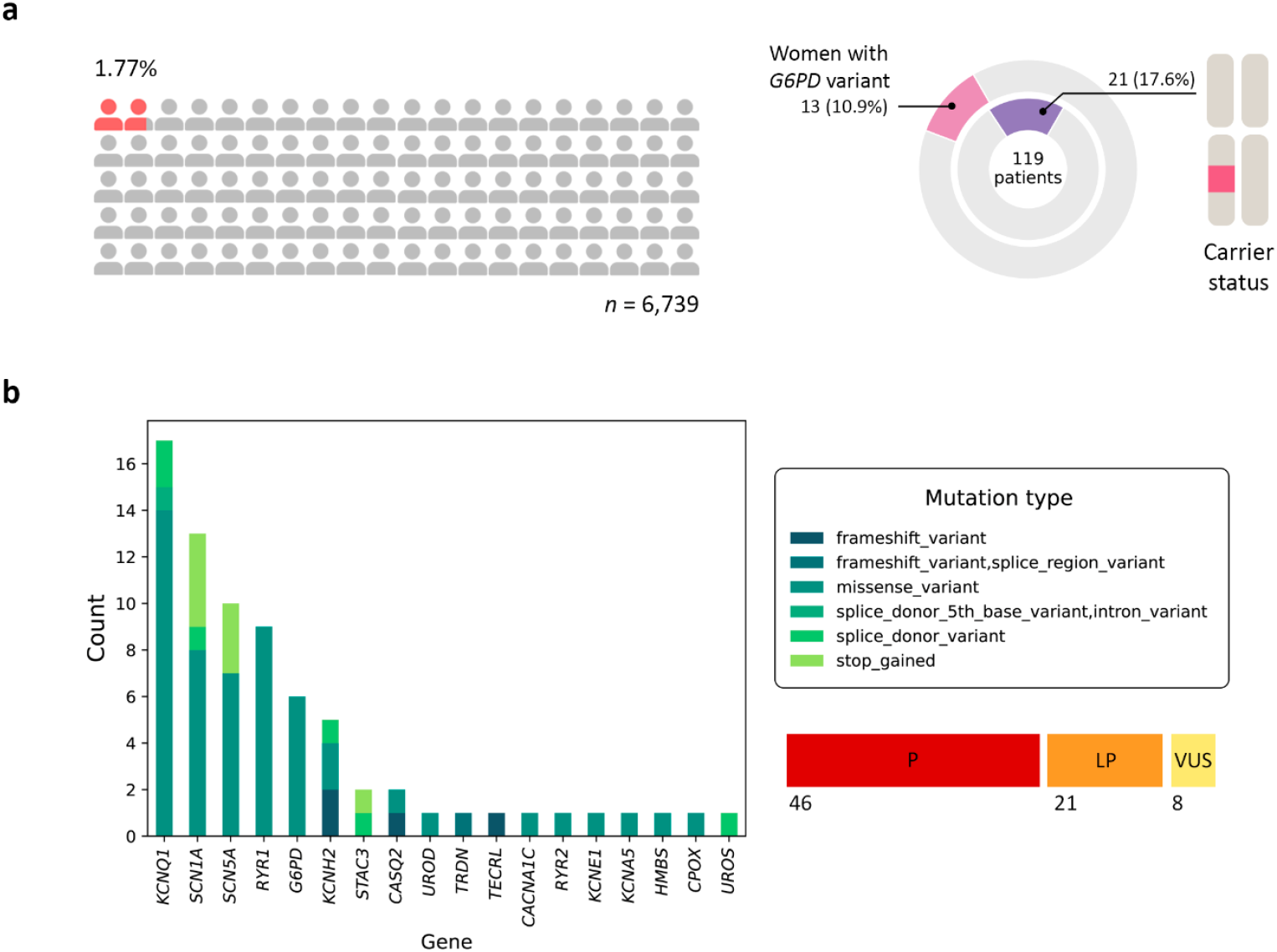
Population distribution and genetic architecture of pharmacogenetically significant monogenic conditions. **A**. Proportion of the screened population carrying clinically significant rare variants associated with monogenic syndromes predisposing to disease (*n* = 119, 1.77% of the total cohort). The distribution of carriers is shown separately: 21 individuals, including 13 women carrying variants in the X-linked *G6PD* gene. **B**. Distribution of unique variants across genes associated with pharmacogenetically significant monogenic disorders: a total of 75 variants, including 46 pathogenic (P), 21 likely pathogenic (LP), and 8 variants of uncertain significance (VUS).

A total of 75 unique genetic variants were identified across 18 genes (Figure 1B, Table S1): *CACNA1C, CASQ2, CPOX, G6PD, HMBS, KCNA5, KCNE1, KCNH2, KCNQ1, RYR1, RYR2, SCN1A, SCN5A, STAC3, TECRL, TRDN, UROD*, and *UROS*. According to ACMG classification, the majority of variants were interpreted as pathogenic (P; 46/75, 61.33%) or likely pathogenic (LP; 21/75, 28%), whereas 8 variants (10.67%) were classified as variants of uncertain significance (VUS). Missense mutations predominated (55 variants, 73.33%), while nonsense and frameshift mutations accounted for 17.33%, and splice-site variants comprised the remaining ∼10% of detected alterations. Notably, we also examined variants located within 10 bp of exon boundaries due to acceptable coverage (>14×), which is an atypical approach for WES; the most distal variant identified was *KCNQ1*(NM_000218.3):c.477+5G>A in a single individual.

### 3.2. Spectrum of pharmacogenetically relevant genetic syndromes

Data on ADRs for long QT syndrome (LQTS) were obtained from the online resource CredibleMeds® (Drugs with Known Risk of TdP, Only Marketed Drugs) [28]; for Brugada syndrome (BrS), from BrugadaDrugs.org [29]; for G6PDD and porphyrias, from the study by Micaglio et al. [10]; and for MH, from CPIC recommendations [30]. ADRs associated with other syndromes were assessed exclusively based on data from the published literature (Table S2).

As shown in Figure 2A, several genes are associated with multiple clinical syndromes, highlighting the complexity of genotype–phenotype relationships underlying pharmacogenetic risk. Among all detected conditions, LQTS was the most frequent, observed in 35 individuals (29.41% of genotype-positive subjects), followed by MH (27 individuals, 22.69%) and G6PDD (20 individuals, 16.81%). Less frequent conditions included DS (DS; 15 individuals, 12.61%), Brugada syndrome (BrS; 8 individuals, 6.72%), porphyrias (6 individuals, 5.04%), catecholaminergic polymorphic ventricular tachycardia (CPVT; 6 individuals, 5.04%), and monogenic atrial fibrillation (AF; 2 individuals, 1.68%) (Figure 2B).

**Figure 2.**
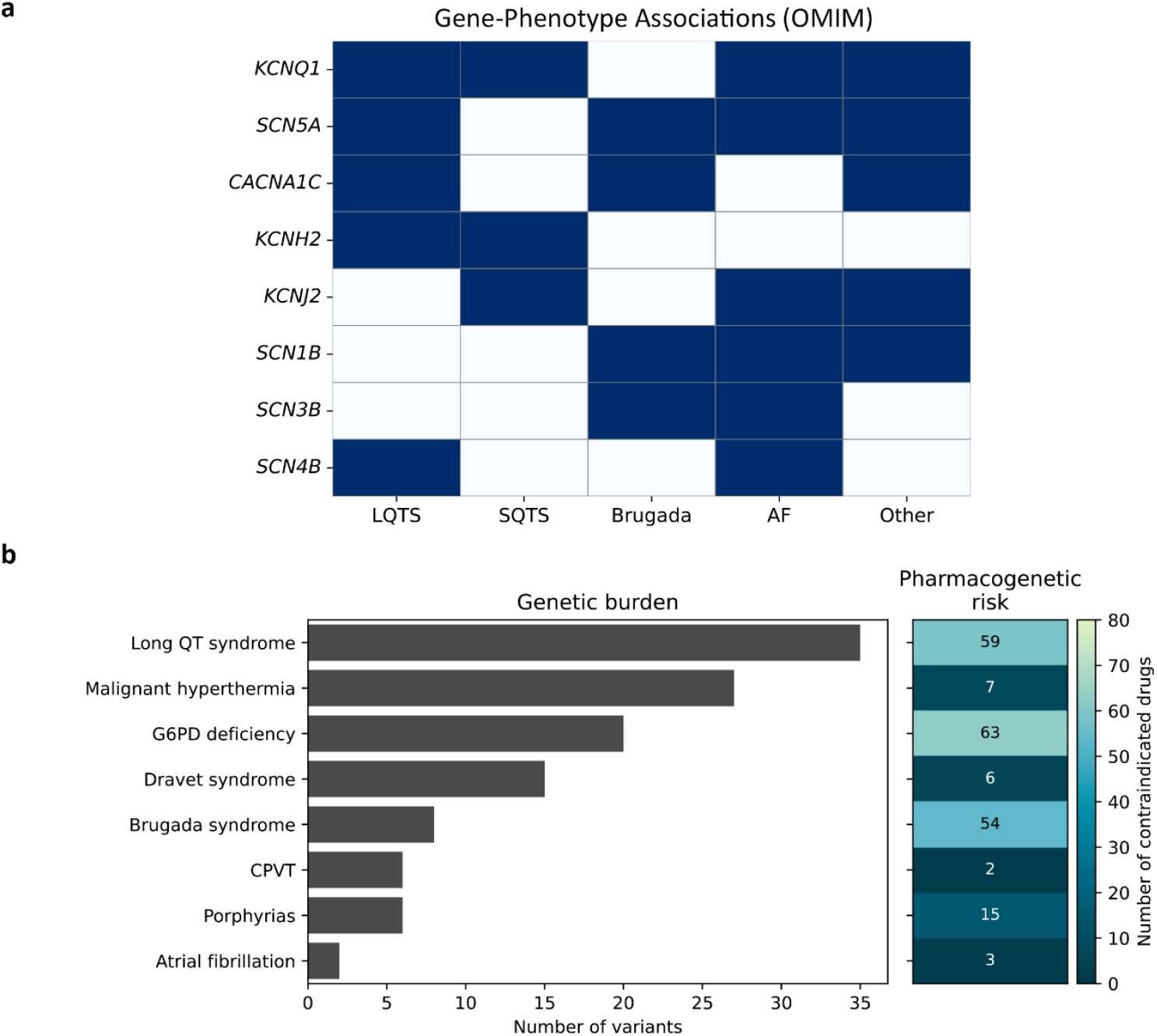
Genetic and clinical heterogeneity of pharmacogenetically significant syndromes. **A**. Overlap of hereditary arrhythmogenic syndromes at the gene level, demonstrating pleiotropy and the potential for a single gene to be associated with multiple phenotypes. **B**. Comparison of the number of patients with syndromes and the corresponding number of drugs associated with an increased risk of adverse drug reactions. Notes: LQTS—long QT syndrome, SQTS—short QT syndrome, AF—atrial fibrillation, CPVT—catecholaminergic polymorphic ventricular tachycardia.

Gene-level analysis revealed *KCNQ1* as the primary contributor to LQTS, harboring the largest number of associated variants (24 variants, 18 unique). *KCNH2* (*n* = 6, 5 unique) and *SCN5A* (*n* = 3) also significantly contributed to cardiac pharmacogenetic risk, reflecting their pleiotropic involvement in multiple arrhythmogenic syndromes. Additionally, single variants with corresponding carriers were detected in *TRDN, CACNA1C*, and *KCNE1. RYR2* was the principal gene underlying CPVT; however, only one carrier was identified, whereas *TECRL, TRDN*, and *CASQ2* variants collectively affected four additional individuals. Variants in *HMBS, UROS, UROD*, and *CPOX* causing porphyria were mostly singular occurrences, although *UROS* (NM_000375.3):c.63+1G>A was observed in three individuals; its clinical significance was lower due to carrier status compared to, for instance, *CPOX*(NM_000097.7):c.991C>T (p.Arg331Trp), which is associated with autosomal-dominant coproporphyria. Among individuals predisposed to MH, *RYR1* variants predominated (9 unique variants; 14 carriers), whereas *STAC3* variants were fewer in number (2 unique) but present in a similar number of carriers (13 individuals). Hemizygous *G6PD* variant carriers accounted for 35%, with no homozygous women identified. BrS in this cohort was associated exclusively with *SCN5A* variants.

Figure 2B summarizes, for each syndrome, both the number of affected patients and the corresponding number of drugs associated with increased ADR risk that should be avoided. Notably, LQTS and BrS were associated with the highest number of contraindicated drugs, emphasizing their particular relevance in the context of preventive pharmacogenomic screening.

## Discussion

Among the 608 entries in the FDA table “Pharmacogenomic Biomarkers in Drug Labeling,” G6PDD (*n* = 41) and MH (*n* = 4) were predominantly represented [31], suggesting that the current pharmacogenetic regulatory framework is insufficiently adapted to account for risks associated with rare monogenic conditions. Nevertheless, out of 48 genes listed, 16 had documented associations in PharmGKB, as accessed via VarSome (retrieved December 13, 2025) [32]; *KCNH2* ranked second after *G6PD* in terms of the number of associated drugs, while some single reported associations did not involve the primary phenotype (e.g., montelukast use in asthma and *ABCC9* variants [33]). Importantly, carriers of variants in these genes may be required to avoid or use with extreme caution at least 179 drugs and other substances spanning multiple pharmacological classes, including analgesics and nonsteroidal anti-inflammatory drugs, antimalarial agents, antimicrobials (antibiotics, antifungals, and antiprotozoals), antiarrhythmic drugs, local and general anesthetics, anticonvulsants, psychotropic medications, diuretics, antidiabetic agents (particularly sulfonylureas), immunosuppressants, prokinetic agents, oncological and hematological therapies, antidotes, antigout medications, sympathomimetic agents, vitamins and vitamin-like compounds, as well as industrial and chemical substances with pharmacological effects. Notably, the studied cohort also comprised individuals harboring single alleles in genes with recessive inheritance, which, while not necessarily associated with overt disease, has important implications for genetic counseling and therapeutic decision-making.

Inherited arrhythmias represent a significant, albeit relatively rare, cause of sudden cardiac death (SCD). In the general population, SCD incidence ranges from approximately 1 per 100,000 in adolescents to 1 per 1,000 among individuals aged 45–75, with a substantial proportion of sudden unexplained deaths in young adults and children (∼10–15%) attributable to primary arrhythmogenic channelopathies in the absence of structural heart disease, such as LQTS, CPVT, and BrS [34]. CPVT is a less commonly recognized syndrome in predictive pharmacogenetics, most frequently caused by *RYR2* variants. It manifests in children and adolescents during physical exertion or emotional stress, despite structurally normal hearts [35]. Its prevalence in Europe is estimated at approximately 1:10,000, and untreated disease carries a high risk of SCD, reaching up to 31% by age 30. A critical clinical feature of CPVT is that standard resuscitative and pharmacologic interventions, particularly catecholamine administration, may exacerbate arrhythmogenicity and lead to fatal outcomes [36].

Another condition often overlooked in pharmacogenomics is AF. GenCC classifies three genes as “Strong” (*KCNQ1, GJA5, MYL4*) and three as “Moderate” (*KCNA5, NPPA, SCN2B*) for monogenic AF. AF is one of the most prevalent arrhythmias, with a lifetime risk of 1:3–5 after age 45 and a global prevalence increase from 33.5 million to 59 million cases between 2010 and 2019 [37]. In our cohort, only *SCN5A*(NM_000335.5):c.1282G>A (p.Glu428Lys) (“Supportive”) and *KCNA5*(NM_002234.4):c.143A>G (p.Glu48Gly) variants were identified in two individuals, underscoring the limited representation of monogenic AF components detectable via WES. Despite the large number of drugs that can precipitate AF, only three are explicitly flagged for patients with pre-existing AF: acetylcholine, adenosine, and ibrutinib [38].

Although the classical neonatal estimate of LQTS prevalence (∼1:2,000) remains widely cited [39], population-based studies in Norway suggest that 1 in 100 individuals carries a pathogenic variant causing LQTS [40]. Limited Russian data also indicate a higher detection rate (1.96% among 2,140 psychiatric patients) [41]. The high population frequency of LQTS-associated variants combined with low clinical penetrance creates a substantial hidden-risk group in which life-threatening arrhythmias may be triggered by routine medications [42]. Notably, in 15–20% of individuals with a phenotypic LQTS presentation, no causative variant is identified; however, their clinical risk is comparable to genotype-positive patients [43]. In our study, this nosological group was the most frequently identified, representing 0.52% of the total cohort. Additionally, no patients with short QT syndrome (SQTS) were observed, likely reflecting the extremely low prevalence of this condition, with only approximately 43 clinically confirmed cases worldwide as of January 2025 [44].

BrS has an estimated global prevalence of approximately 1 in 2,000 individuals, comparable to that of LQTS, with substantially higher rates reported in Southeast Asia—up to 35.5 per 1,000 overall and up to 17.7 per 1,000 in Thailand [45]. Although associations with BrS have been proposed for 22 genes to date, only *SCN1B* and *SCN5A* have achieved a “Definitive” level of evidence according to GenCC [46]. In BrS cohorts published between 2002 and 2022, genetic testing was performed in an average of 59% of patients, with an overall diagnostic yield of 26%. Both testing rates and diagnostic yield varied markedly across regions, ranging from 10% in Hong Kong to 100% in selected centers in China, Japan, France, and Spain, while diagnostic yield ranged from 10% in Japan to 67% in Belgium [47].

At the same time, it is important to distinguish true Brugada syndrome from the Brugada ECG pattern, which may occur transiently and be induced by fever, medications, or electrolyte disturbances, without clinical manifestations of the disease. The Brugada pattern is considerably more common than overt BrS and is often detected in individuals without a family history of sudden cardiac death, substantially complicating the interpretation of genetic findings and clinical risk assessment [48]. For example, a Brugada ECG pattern has been described in a patient with hypertrophic cardiomyopathy carrying pathogenic variants in *MYBP3* and *MYH7* [49]. Contemporary clinical observations and experimental data further indicate a close relationship between Brugada syndrome and arrhythmogenic right ventricular cardiomyopathy/dysplasia (ARVC/D). In some patients, features of BrS and ARVC/D may coexist, combining characteristic Brugada ECG patterns with structural abnormalities of the right ventricle that meet diagnostic criteria for ARVC/D. In our study, no clinically significant *PKP2* variants were identified; however, this gene was included based on its reported overlap with ARVC/D type 9 and its “Limited” evidence of association with BrS according to GenCC [50,51].

Variants in *RYR1* are identified in the majority of patients with MH, accounting for approximately 50–60% of cases. Notably, pathogenic *RYR1* variants are far more common in the general population (approximately 1 in 800 individuals), whereas a clinical MH crisis occurs in only 1 per 10,000–150,000 anesthetic exposures. Nevertheless, in the absence of timely treatment, MH-related mortality may reach up to 80% [52]. In our cohort, the number of carriers of *RYR1* and *STAC3* variants was approximately comparable when accounting for carrier status in all *STAC3*-positive individuals; however, the number of unique *RYR1* variants exceeded that of *STAC3* by a factor of 4.5. Patients with *STAC3* variants frequently require surgical interventions due to multiple congenital anomalies, necessitating heightened vigilance during anesthesia. Given the autosomal recessive inheritance pattern, parents of affected individuals may be asymptomatic carriers [53]. The prevalence of *STAC3*-associated MH is difficult to estimate but appears to be extremely low based on the limited available literature [54].

DS is a rare epileptic encephalopathy with an incidence of approximately 2.2–6.5 cases per 100,000 individuals and a prevalence of 1.2–6.5 per 100,000 population (up to 0.0065%). In our cohort, the prevalence of genotype-positive DS was 0.22%, exceeding global estimates. Despite its rarity, DS is associated with high mortality (approximately 15.8 per 1,000 patient-years, primarily due to sudden unexpected death in epilepsy [SUDEP]) and an earlier age at death [55]. It is well established that sodium channel blockers, particularly carbamazepine, oxcarbazepine, and lamotrigine, may exacerbate seizures and worsen cognitive outcomes in DS. Other antiepileptic drugs, including vigabatrin, rufinamide, and phenobarbital, may also increase seizure frequency in some patients [56]. The economic burden of DS is substantial, with costs estimated to be 1.5-fold higher than those for drug-resistant epilepsy and fivefold higher than for epilepsy in remission; mean annual costs have reached up to €29,872 in a multicenter German study [57].

Porphyrias constitute a group of rare disorders with prevalence varying widely by clinical subtype. In Europe, the most common forms—porphyria cutanea tarda, acute intermittent porphyria, and erythropoietic protoporphyria—occur at approximate frequencies of 1:10,000, 1:20,000, and 1:50,000–75,000, respectively. A decline in symptomatic acute hepatic porphyrias alongside increased detection of erythropoietic protoporphyria has been observed, likely reflecting improved diagnostics and reduced exposure to triggering factors [58]. In children, porphyrias often mimic common pediatric conditions, and clinical manifestations are frequently precipitated by medications and other external factors. Certain antiepileptic drugs (e.g., carbamazepine, phenytoin, valproate) are known to worsen disease severity. Given the overlap of clinical and biochemical features among porphyria subtypes, molecular confirmation is crucial for selecting safe therapies and identifying asymptomatic carriers within families [59].

G6PDD remains a major global public health concern. By 2021, its prevalence had reached 443 million cases worldwide, representing an increase of more than 80% compared with 1990, with the greatest burden observed in South Asia. The condition is more common in males, particularly in childhood and older age, and exhibits pronounced regional variability, underscoring the need for expanded screening programs and targeted public health interventions, especially in low- and middle-income countries [60]. In our cohort, G6PDD ranked third in frequency, with the important consideration that 13 women were identified as carriers.

## Limitation of Our Study

We focused on variants previously reported as P or LP in ClinVar, excluding those annotated as benign, which likely led to an underestimation of the true population prevalence of causative variants. This is particularly relevant for variants with high ACMG classification scores that have not yet been described in clinical practice. It should be noted that LOF variants predicted to be disease-causing, even when identified for the first time in a given individual, are important for personalized therapy and for assessing the risk of severe ADRs, although they require further functional and clinical validation. In addition, we did not include genes with insufficiently robust evidence for disease association, even in cases where convincing animal models have been established, such as *ZFHX3* in the context of AF [61].

The presence of a pathogenic variant does not invariably translate into a clinical phenotype: some identified variants may exhibit incomplete penetrance or require additional genetic or environmental factors for disease manifestation, as observed in susceptibility to MH. This limits the direct translation of carrier status into an absolute probability of ADRs but does not exclude an increased risk.

Although our cohort was relatively large (*n* = 6,739), it may not fully capture the ethnic and regional diversity of the Russian population; therefore, the observed frequencies require validation in independent cohorts.

Finally, WES has inherent limitations in detecting copy number variants and noncoding regions that may contribute to ADR risk, as well as other methodological constraints typical of next-generation sequencing approaches.

## Conclusions

For the first time in a large cohort of 6,739 individuals from the Russian population, we demonstrate that at least 1.77% carry genetic variants that substantially increase the risk of severe ADRs associated with commonly used medications. LQTS syndrome and inherited arrhythmias were the most frequently identified conditions, which, in the absence of adequate clinical awareness, may lead to sudden cardiac death, followed by MH, which is of particular relevance in the surgical and anesthetic setting. Incorporation of extended analyses of monogenic syndromes into standard pharmacogenomic testing panels has the potential to significantly improve the safety of pharmacotherapy.

## Supporting information

A total of 75 unique genetic variants were identified across 18 genes (Figure 1B, Table S1)

ADRs associated with other syndromes were assessed exclusively based on data from the published literature (Table S2).

## Supplementary Material

Table S1. List of identified genetic variants. Table S2. Syndrome-specific adverse drug reactions relevant for clinical practice.

## Institutional Review Board Statement

This study was conducted in accordance with the Declaration of Helsinki, and approval was obtained from the Local Research Ethics Committee of Russian National Medical University (Protocol No. 241, from 26 June 2024), and all participants provided written informed consent prior to data collection.

## Author Contributions

Buianova A.A. – formal analysis, conceptualization, methodology, writing— original draft preparation; Cheranev V.V., Kuznetsov M. Iu – data curation, software; Shmitko A.O., Ilyina G.A., Belova V.A. – investigation; Vasiliadis Iu.A., Suchalko O.N. – software; Dmitriy O. Korostin – resources, supervision, writing—review and editing.

## Informed Consent Statement

Informed consent was obtained from all subjects involved in this study.

## Data Availability Statement

The sequence data are generated from patient samples and therefore are only available under restricted access.

## Conflicts of Interest

The authors declare no conflicts of interest. The funders had no role in the design of the study; in the collection, analyses, or interpretation of data; in the writing of the manuscript; or in the decision to publish the results.

## Notes

### Competing Interest Statement

The authors have declared no competing interest.

### Funding Statement

This study was funded.

